# Maternal and congenital syphilis at a national referral center in Peru, 2023-2025

**DOI:** 10.64898/2026.03.04.26347675

**Authors:** William Leung, Carlos Velásquez Vásquez, Luis Meza Santivañez, Pedro Arango-Ochante, Kelika A. Konda, Silver K. Vargas, Carlos F. Cáceres, Jeffrey D. Klausner, Lao-Tzu Allan-Blitz

## Abstract

**Background:** Congenital syphilis remains a preventable cause of neonatal morbidity and mortality despite the availability of inexpensive diagnostics and effective treatment. We aimed to evaluate the maternal and congenital syphilis burden at the Instituto Nacional Materno Perinatal, Peru’s national referral center for maternal and perinatal care.

**Methods:** We conducted a retrospective analysis of aggregated, de-identified surveillance data from January 2023 to December 2025. We calculated the prevalence of maternal syphilis and incidence of congenital syphilis, and compared those estimates with World Health Organization (WHO) elimination benchmarks.

**Results:** Among 59,598 pregnant women screened, maternal syphilis prevalence ranged between 1.0% and 1.2%. Across 36,094 live births, congenital syphilis incidence ranged between 383 and 526 per 100,000 live births, consistently exceeding the WHO target of 50 per 100,000 live births. Over half of maternal infections were diagnosed at outside facilities before referral. Reported treatment coverage exceeded 90% among mothers and reached 100% among infants.

**Conclusions:** Congenital syphilis incidence consistently exceeded elimination benchmarks despite relatively stable maternal syphilis prevalence, highlighting potential gaps in timely diagnosis and linkage between diagnosis and treatment.

## Introduction

*Treponema (T.) pallidum,* the infectious cause of syphilis, affected more than seven million adults globally in 2020.^1^ Untreated *T. pallidum* infection during pregnancy can lead to congenital syphilis, which is a major cause of stillbirth and miscarriage. Among affected infants, congenital syphilis can result in anemia, hepatobiliary dysfunction, mucocutaneous lesions, and if untreated, serious neurological and musculoskeletal complications or death.^2^ The consequences of congenital syphilis extend beyond the infant and family. Mother-to-child transmission of syphilis contributes to $437 million in additional medical spending worldwide (adjusted to 2025 USD).^3^

Effective, low-cost diagnostics and treatment have been available for decades. Inexpensive, point-of-care syphilis tests are widely available and enable same-day therapy.^4–6^ Treatment of syphilis during pregnancy prevents more than 98% of congenital syphilis cases.^7^ Additionally, two separate studies estimated that the cost per pregnant woman screened was less than $3 USD.^8,9^ Consequently, screening for syphilis during pregnancy is among the most cost-effective reproductive health interventions, with many regulatory bodies now recommending universal third trimester screening for syphilis.^10–12^ Yet, more than half of congenital syphilis cases globally occur among women who were never screened.^13^

Another consequence of limited antenatal screening is a paucity of epidemiologic data describing the burden of congenital syphilis in low- and middle-income countries. As of 2017, among 81 countries whose data were collected under the WHO’s “Countdown” program, only 53 reported any congenital syphilis surveillance indicators, and only 41 reported syphilis treatment coverage.^14^ Across Latin America, many countries continue to report an incidence of congenital syphilis above the WHO elimination threshold of 50 cases per 100,000 live births,^13^ ranging from 129 cases per 100,000 live births in Mexico in 2016, to 197 cases per 100,000 live births in Paraguay in 2018.^15,16^ A 2019 national estimate in Peru reported 95 cases of congenital syphilis per 100,000 live births.^16^ While national estimates can provide important benchmarks, they often fail to provide actionable insight into operational gaps along the continuum of care that may be apparent within data from high-volume referral centers. This present study aimed to characterize the burden of maternal and congenital syphilis at the Instituto Nacional Materno Perinatal in Lima, Peru.

## Methods

### Study setting

The Instituto Nacional Materno Perinatal in Lima serves as Peru’s national referral center for maternal and perinatal care. In addition to providing medical services to women, the center maintains a key role in the surveillance and management of sexually transmitted infections during pregnancy. It is categorized as a Level III-2 health facility, the highest tier in Peru’s national health system and indicative of specialized, high-complexity care, with 370 beds as of 2024. Although it serves patients from across Peru, more than 90% of admissions come from Metropolitan Lima.^17^ Specifically, 37% of patients are from San Juan de Lurigancho, the most populous and among the most socio-economically marginalized districts in the region.^18^

### Data elements and analysis

We retrospectively analyzed aggregate surveillance data from the Instituto Nacional Materno Perinatal between January 2023 and December 2025. We limited the study to those years as they were the data that were most complete and standardized.

At the Instituto Nacional Materno Perinatal, clinicians routinely screen all pregnant women presenting to the Emergency and Preventive Medicine services department, at all stages of pregnancy, for syphilis using an initial treponemal rapid test, followed by lipoidal (non-treponemal) testing with rapid plasma reagin (RPR) (HUMAN diagnostics, Germany). For patients with reactive initial treponemal results, clinicians then perform confirmatory treponemal hemagglutination assay (TPHA) using a TPHA Cromatest (Linear Chemicals, Spain). Peru’s National Resource Supply Center serves as the distributor of both tests.

We classified maternal cases as active infection when RPR titers were at least 1:4 dilutions with a positive rapid syphilis test and TPHA. We classified cases as congenital syphilis if a) the child was born to a mother with untreated or inadequately treated syphilis, defined as failure to complete a penicillin-based regimen appropriate for the stage of infection that was initiated 30 or more days before delivery, b) the infant’s RPR titer was at least two-times greater than that of the mother, or c) the child presented with symptoms consistent with congenital syphilis; or d) the pregnancy resulted in miscarriage or neonatal death attributable to congenital syphilis.^19^ We classified cases as exposed if infants were born to mothers with confirmed active syphilis during pregnancy, but who did not meet the above criteria for congenital syphilis.

We extracted epidemiologic data from the Instituto Nacional Materno Perinatal’s internal epidemiologic surveillance office. The maternal variables included in the analysis were maternal age and whether initial diagnosis was at or outside the Instituto Nacional Materno Perinatal. Perinatal outcomes were classified as exposed newborn, congenital syphilis, miscarriage due to congenital syphilis, or neonatal death due to congenital syphilis.

We calculated the prevalence of maternal syphilis, congenital syphilis incidence per 100,000 live births, and treatment coverage among mothers and neonates, and compared these metrics to international elimination benchmarks set by the WHO.^16^ We aggregated data annually to describe temporal trends in screening, diagnosis, and outcomes for 2023, 2024, and 2025. We used descriptive statistics to summarize frequencies and proportions of all covariates.

### Ethical considerations

The study was approved by the Institutional Research Ethics Committee of the Instituto Nacional Materno Perinatal (Approval No. 020-2026-CIEI/INMP). The study used de-identified surveillance data and did not involve patient contact or intervention.

## Results

Between January 2023 and December 2025, the Instituto Nacional Materno Perinatal screened a total of 59,598 pregnant women for syphilis, with 19,657 women screened in 2023, 20,906 in 2024, and 19,035 in 2025 (Table 1). In 2023, 324 pregnant women tested positive on the initial treponemal rapid test, and confirmatory testing confirmed active syphilis in pregnancy by combined rapid test, RPR, and TPHA reactivity in 192 women (1.0% of those screened). The number of confirmed active infections was 242 (1.2%) in 2024, and 193 (1.0%) in 2025. Across all years, more than half of all confirmed cases of active syphilis had been diagnosed outside of the Instituto Nacional Materno Perinatal prior to referral – 54% in 2023, 67% in 2024, and 65% in 2025. Adolescents (<19 years) represented 15% of confirmed active cases in 2023, decreasing to 7.0% in 2024 and 15% in 2025. Pregnant women older than 35 years accounted for 10%, 5.8%, and 11% of cases across the respective periods.

**Table 1:** Maternal outcomes of all women screened with treponemal rapid test at the Instituto Nacional Materno Perinatal.

|  | **2023** | **2024** | **2025** |
| --- | --- | --- | --- |
| Number screened with rapid test | 19,657 | 20,906 | 19,035 |
| Women with reactive test (including memory cases) | 324 (1.6%) | 371 (1.8%) | 362 (1.9%) |
| Confirmed active cases | 192 (1.0 % of overall screened) | 242 (1.2% of overall screened) | 193 (1.0% of overall screened) |
| Diagnosed outside the Instituto Nacional Materno Perinatal | 104 (54% of confirmed active cases) | 163 (67% of confirmed active cases) | 126 (65% of confirmed active cases) |
| Diagnosed within the Instituto Nacional Materno Perinatal | 88 (46% of confirmed active cases) | 79 (33% of confirmed active cases) | 67 (35% of confirmed active cases) |
| Adolescents (<19 years) | 28 (15% of confirmed active cases) | 17 (7.0% of confirmed active cases) | 29 (15% of confirmed active cases) |
| >35 years | 20 (10% of confirmed active cases) | 14 (5.8% of confirmed active cases) | 21 (11% of confirmed active cases) |

Regarding perinatal outcomes, the Instituto Nacional Materno Perinatal recorded 12,249 live births in 2023, 12,349 in 2024, and 11,496 in 2025 (Table 2). Among those, clinicians classified 129 (1.1%), 147 (1.2%), and 101 (0.9%) newborns as exposed to maternal syphilis, respectively. Across the included years, the Instituto Nacional Materno Perinatal identified 55 congenital syphilis cases in 2023, 65 in 2024, and 44 in 2025, corresponding to incidences of 449, 526, and 383 per 100,000 live births, respectively. Among these congenital syphilis cases, 22, 32, and 22 resulted in miscarriage, and five, one, and zero resulted in neonatal death in 2023, 2024, and 2025, respectively.

**Table 2:** Neonatal outcomes of all live births at the Instituto Nacional Materno Perinatal.

|  | 2023 | 2024 | 2025 |
| --- | --- | --- | --- |
| Total live births in the Instituto Nacional Materno Perinatal | 12,249 | 12,349 | 11,496 |
| Exposed newborns | 129 (1.1% of total live births) | 147 (1.2% of total live births) | 101 (0.9% of total live births) |
| Liveborn with congenital syphilis | 28 (0.2% of total live births) | 32 (0.3% of total live births) | 22 (0.2% of total live births) |
| Miscarriage due to congenital syphilis | 22 (0.2% of total live births) | 32 (0.3% of total live births) | 22 (0.2% of total live births) |
| Neonatal death due to congenital syphilis | 5 (0.04% of total live births) | 1 (0.01% of total live births) | 0 |

**Table 3:** Key indicators on maternal syphilis prevalence and congenital syphilis incidence, and maternal and neonatal treatment coverage compared to WHO global target benchmarks.

|  | **WHO Global**<br>**Target** | **2023** | **2024** | **2025** |
| --- | --- | --- | --- | --- |
| Maternal prevalence | 0.7%<br>(estimated global burden) | 1.0% | 1.2% | 1.0% |
| Congenital syphilis incidence | 50 per 100,000 live births | 449 per 100,000 live births | 526 per 100,000 live births | 383 per 100,000 live births |
| Maternal treatment coverage | 95% | 91% | 95% | 91% |
| Neonatal treatment coverage | 100% | 100% | 100% | 100% |

In 2023, 91% of women and 100% of neonates completed treatment. In 2024, 95% of women and 100% of neonates completed treatment. And in 2025, 91% of women and 100% of neonates completed treatment.

## Discussion

The present analysis found that maternal syphilis prevalence at the Instituto Nacional Materno Perinatal remained relatively stable between 2023 and 2025, ranging between 1.0% and 1.2%, and only slightly exceeding the 2016 global estimate of 0.7% among pregnant women. In contrast, congenital syphilis incidence ranged from 383 to 526 per 100,000 live births, consistently surpassing the WHO’s elimination benchmark of <50 per 100,000 live births and significantly higher than the national estimate of 95 per 100,000 live births in 2019.^16,20^ Across all years, treatment metrics largely aligned with WHO elimination targets of 95% treatment coverage among mothers and 100% coverage among infants.^21^

These findings suggest that despite a maternal prevalence similar to prior global estimates, the observed congenital syphilis incidence was substantially elevated, indicating a potential discordance between the magnitude of congenital disease and the relative stability of maternal infection burden. As Peru’s national referral center for maternal and perinatal care, the Instituto Nacional Materno Perinatal manages a complex patient population; nonetheless, referral status alone may not fully explain the observed difference between maternal syphilis prevalence and congenital syphilis incidence relative to national and global benchmarks.

Persistent congenital syphilis incidence despite the availability of effective diagnostics and treatment suggests that opportunities for prevention continue to be missed. Because surveillance data were available only in aggregate form, individual-level linkage between maternal and infant records was not possible. As a result, the number of reported maternal infections within a given year did not necessarily correspond to the total number of reported neonatal outcomes, and thus this present analysis was not designed to identify where along the prevention cascade congenital infections occurred. Nonetheless, prior literature points towards four key gaps. First, many women are not identified as pregnant or do not present for antenatal care until late in gestation. Even a small fraction of women with missed or late-detected/late-treated syphilis carries a high vertical transmission risk, estimated at 50-90% depending on disease stage; further, 94% of congenital syphilis-related adverse outcomes occur in women who do not receive timely and adequate screening and treatment during pregnancy.^22,23^ Second, gaps in screening implementation may arise from health system barriers. Those include inconsistent adherence to screening guidelines, stockouts of diagnostics, insufficient quality management, and limited availability of trained healthcare personnel.^24^ Third, patient- and community-level barriers can disrupt continuity of care and prevent consistent follow-up. Those barriers include financial constraints, geographic distance to health facilities, and poor health literacy. Finally, even when infections are detected, inadequate linkage from screening to treatment can arise due to low provider awareness, particularly for providers who rarely treat cases of syphilis, or via medication stockouts. Early testing and uninterrupted treatment during initial stages of pregnancy remain the decisive factors in preventing congenital syphilis.

In this study, more than half of the women included had been initially diagnosed with syphilis at outside facilities before referral to the Instituto Nacional Materno Perinatal. The observed increase in this proportion over time could indicate that community-level screening programs have successfully identified cases of syphilis. However, the incidence of congenital syphilis might suggest that detection alone may not be translating into effective prevention. That underscores the importance of the linkage between screening and timely, adequate treatment. Some referred women may have begun treatment late, received inadequate or interrupted treatment prior to arrival at the Instituto Nacional Materno Perinatal, or experienced reinfection from untreated partners. However, those data were not available for analysis. A recent study at the Instituto Nacional Materno Perinatal reported the persistence of congenital syphilis in infants whose mothers received proper treatment for syphilis, with the aforementioned factors cited as the most likely reasons.^22^

Point-of-care syphilis tests directly address several of the upstream challenges, including delayed diagnosis by enabling same-day testing and treatment initiation at the first antenatal encounter. Implementation studies at the Instituto Nacional Materno Perinatal and a network of 15 sub-urban community health centers across Lima have shown that adoption of rapid syphilis testing substantially reduced the number of required clinic visits and improved the screening-to-treatment cascade.^25^ Beyond operational efficiency, multiple modelling studies have also demonstrated that rapid syphilis testing, particularly when integrated with rapid HIV testing, is cost-saving across multiple low- and middle-income country settings.^26,27^ Point-of-care testing alone, however, does not address all barriers to follow-up, timely treatment completion, partner management, or reinfection.

Globally, the challenges observed at the Instituto Nacional Materno Perinatal reflect the broader difficulty faced by many low-resource settings in congenital syphilis elimination. In 2007, the WHO launched a global initiative to eliminate congenital syphilis as a public health threat by 2030, defining the impact target as <50 cases per 100,000 live births.^13^ Over the past decade, the WHO further established process targets to achieve this goal.^21^ Those include 95% of pregnant women receiving antenatal care with timely syphilis testing, and 95% of women who test positive receiving adequate treatment. Although Peru has committed to similar elimination targets, the findings from this study indicate continued efforts are needed to continue progress towards elimination targets.

## Limitations

This study had several limitations. First, the study relied on aggregated surveillance data from a single national referral center, limiting generalizability to other regions of the country and to non-referral settings. Similarly, because the surveillance dataset was only available in aggregate form, we were unable to perform individual-level linkage between maternal and infant records. Moreover, the surveillance data did not include information regarding partner treatment, timing of maternal treatment, or other key elements of the congenital syphilis prevention cascade, limiting our ability to pinpoint specific drivers of persistent transmission. Finally, given the late manifestation of some congenital syphilis cases, which can occur up to two years post-birth, it is likely that this study underreports the true congenital syphilis incidence within the sample population.^2^

## Conclusion

Congenital syphilis incidence between 2023 and 2025 consistently exceeded national and WHO elimination targets in Peru’s national referral center for maternal and perinatal care. Further investigation with individual-level data is needed to identify specific upstream factors contributing to persistent congenital transmission and to inform targeted interventions that contribute to reaching congenital syphilis elimination goals.

## Acknowledgements

We thank all the staff at the Emergency and Preventive Medicine services department of the Instituto Nacional Materno Perinatal.

## Statements and declarations

### Author contributions

WL conducted the primary data analysis, wrote the initial draft of the manuscript, and contributed to revisions. CVV, LMS, and PA-O supported data curation and interpretation, and contributed to revisions of the manuscript. KAK, SKV, CFC, and JDK, contributed to data interpretation, project oversight, and revisions of the manuscript. LT-AB conceived the project, assisted with data curation, analysis and interpretation, obtained funding, contributed to revisions of the manuscript and provided project oversight. All authors confirm that they had full access to all the data in the study and accept responsibility to submit for publication.

### Consent to participate

Not applicable.

### Consent for publication

Not applicable.

### Data availability

The aggregated de-identified data supporting the findings of this study are available upon request from the corresponding author with publication.

